# Evaluating the effects of a community-based Pre-Conception Care package through trained nurses on Pre-conception Health, Maternal and Child Health Outcomes: Protocol of a Stepped-wedge Cluster Randomized Implementation Trial

**DOI:** 10.1101/2025.03.28.25324848

**Authors:** Sophiya Kalai, Ashwini Kalantri, Swati Misra, Anuj Mundra, Rutuja Kolhe, Arjunkumar Jakasania, Kamlesh Mahajan, Harshal Sathe, Bharat Patil, Abhishek V. Raut, Chetna Maliye, Shuchi Jain, Satish Kumar, R J Paradkar, Subodh S. Gupta, Poonam Varma, B S Garg

**Affiliations:** Department of Community Medicine, Mahatma Gandhi Institute of Medical Sciences, Sewagram, Wardha, India – 442102; National Health Systems Resource Centre, Delhi, India - 110067; Department of Psychiatry, Mahatma Gandhi Institute of Medical Sciences, Sewagram, Wardha, India – 442102; Department of Pathology, Mahatma Gandhi Institute of Medical Sciences, Sewagram, Wardha, India – 442102; Department of Obstetrics & Gynecology, Mahatma Gandhi Institute of Medical Sciences, Sevagram, Maharashtra, India-442102; Department of Biochemistry, Mahatma Gandhi Institute of Medical Sciences, Sevagram, Maharashtra, India-442102; District Health Office, Zilla Parishad, Wardha, India

**Keywords:** Preconception care, Anemia, Undernutrition, Maternal depression, Stunting, Implementation research, Task shifting, Maternal morbidities, community-based participatory research, Health promotion

## Abstract

**Objectives:** The study aimed to evaluate the impact of village-level preconception care (PCC) delivered by trained ANMs on preconception health and maternal-child health outcomes among eligible women in rural Wardha, Maharashtra, India. It also sought to identify factors influencing the implementation and utilization of the PCC package, assess the feasibility of the intervention using the RE-AIM framework, and analyze the unit cost of delivering community-based PCC interventions.

**Methods:** **Design:** Open cohort stepped-wedge cluster randomized design (swRCT) with repeated cross-sectional sampling over three years, allowing phased implementation of the PCC package and systematic evaluation. **Setting:** Conducted in rural villages of Wardha Community Development block, integrated with the District Health System, using Village Council. **Participants:** Villages under four PHCs agreeing to Social Franchisee agreements were included. Eligible participants were women aged 19–45 years who had not completed their families, consented to participate, and resided in the study area for two years. **Intervention &Comparator:** The PCC package, delivered via village clinics, addressed nutrition, reproductive health, vaccination, and mental health, compared to routine government health services during the control period. **Measurements:** Data collection included baseline demographics, health behaviors (diet, physical activity, stress mediation), anthropometry, hemoglobin levels, and intervention compliance. Analysis used GLM regression models, with subgroup analysis on compliance and outcomes.

**Discussion:** The PCC package, delivered by ANMs in rural Wardha, addresses undernutrition, anemia, and maternal depression. The implementation outcomes.

**Results and Conclusions:** These will be assessed using the RE-AIM (Reach, Effectiveness, Adoption, Implementation, Maintenance) framework, with the results anticipated to be published by the end of 2026, showcasing improved maternal-child health and the feasibility of community-based interventions in rural settings.

**Trial registration:** CTRI/2024/03/064582 [Registered on: 21/03/2024] - Trial Registered Prospectively

## 1. Background

Every person has the right to health, and every newborn deserves the chance to survive and thrive. Preconception care (PCC) differs from prenatal care, as early visits may be too late for interventions. PCC enables timely action before conception, promoting healthier pregnancies and better outcomes. Between 1990 and 2015, maternal mortality fell by 45% and child mortality by over 50%, but many countries, including India, did not fully meet the Millennium Development Goals (MDGs) 4 and 5. (UNITED NATIONS, 2015; United Nations Economic and Social Commission for Asia and Pacific (UNESCAP), 2015) Although these goals sought to reduce child mortality and improve maternal health, progress was hindered by inequalities, resource limitations, and preventable deaths, leaving many low- and middle-income countries short of targets. (United Nations, 2019) Despite effective interventions and increased public health efforts, 15,000 children and 800 women still die daily from preventable or treatable causes. (SUZUKI & KASHIWASE, 2019) Despite improved access to maternal and child health services, 40% of pregnancies are unplanned, leading to unsafe abortions. Maternal undernutrition and iron-deficiency anemia cause 20% of maternal deaths, while teenage pregnancies raise low birth weight and perinatal deaths by over 50%, posing long-term risks. (Bearak et al., 2018; Daru et al., 2018; USAID, 2017; World Health Organization, 2020) Over 90% of maternal and child health challenges are in South Asia and Sub-Saharan Africa’s low- and middle-income countries, where limited resources hinder life-saving interventions. (Gülmezoglu et al., 2016; Lassi et al., 2016)

PCC is vital for improving maternal and child health. Investing in PCC reduces mortality by promoting contraceptive use, preventing unintended pregnancies, and minimizing complications in pregnancy and childbirth. It helps prevent birth defects, preterm births, low birth weight, stillbirths, and neonatal infections, while supporting better antenatal care, breastfeeding, and preventing HIV/STI transmission. PCC addresses undernutrition, risky behaviors like smoking and alcohol, and lowers long-term risks of type 2 diabetes and cardiovascular disease, making it a key global health strategy. Interventions at various life stages, especially before conception, improve maternal and child health by addressing nutritional deficiencies, managing medical conditions, and promoting healthy lifestyle choices, reducing complications and enhancing outcomes across the care continuum (Say et al., 2014; Schoenaker et al., 2024). WHO recommends prenatal care programmes to be implemented in all countries, for promoting good health, prevent health problems and respond to them if they occur during pregnancy (Schoenaker et al., 2024). WHO has included Nutritional conditions, Tobacco use, Genetic conditions, Environmental health, Infertility/sub-fertility, Interpersonal violence, Too-early unwanted and rapid successive pregnancies, Sexually transmitted infections, HIV, Mental health, Psychoactive substance use, Vaccine-preventable diseases and Female genital mutilation as 13 broad areas to be addressed by the PCC package (Ministry of Health and Family Welfare (MOHFW). Government of India, 2015; National Rural Health Mission, 2013). WHO recommends prenatal care programs globally to promote health, prevent issues, and address complications during pregnancy (Schoenaker et al., 2024).

WHO has included Nutritional conditions, Tobacco use, Genetic conditions, Environmental health, Infertility/sub-fertility, Interpersonal violence, Too-early unwanted and rapid successive pregnancies, sexually transmitted infections, HIV, Mental health, Psychoactive substance use, Vaccine-preventable diseases and Female genital mutilation as 13 broad areas to be addressed by the PCC package. WHO has included Nutritional conditions, Tobacco use, Genetic conditions, Environmental health, Infertility/sub-fertility, Interpersonal violence, Too-early unwanted and rapid successive pregnancies, sexually transmitted infections, HIV, Mental health, Psychoactive substance use, Vaccine-preventable diseases and Female genital mutilation as 13 broad areas to be addressed by the PCC package. WHO recommends prenatal care programmes to be implemented in all countries, for promoting good health, prevent health problems and respond to them if they occur during pregnancy. WHO has included Nutritional conditions, Tobacco use, Genetic conditions, Environmental health, Infertility/sub-fertility, Interpersonal violence, Too-early unwanted and rapid successive pregnancies, sexually transmitted infections, HIV, Mental health, Psychoactive substance use, Vaccine-preventable diseases and Female genital mutilation as 13 broad areas to be addressed by the PCC package (Kolhe, 2023; Ministry of Health and Family Welfare (MOHFW). Government of India, 2015; Schoenaker et al., 2024).

The WHO recommends addressing 13 key areas in the PCC package, including nutrition, unintended and rapid pregnancies, STIs (including HIV), infertility, vaccine-preventable diseases, genetic conditions, substance use, environmental health, interpersonal violence, mental health, and female genital mutilation. WHO recommends prenatal care programmes to be implemented in all countries, for promoting good health, prevent health problems and respond to them if they occur during pregnancy. WHO recommends prenatal care programmes to be implemented in all countries, for promoting good health, prevent health problems and respond to them if they occur during pregnancy (Kolhe, 2023; World Health Organization Headquarters, 2012).

In this study, the primary objective is to assess the pre-conception health status among women from eligible couples, focusing on the prevalence of undernutrition (pre-pregnancy BMI <20 kg/m²), anemia (hemoglobin level <12 gm/dl), depression (WHO SRQ-20 score <11), and contraceptive prevalence. Secondary objectives include evaluating maternal health outcomes including age at first conception, the delay of first pregnancy after marriage, birth intervals between pregnancies, and the prevalence of conditions such as pre-eclampsia, eclampsia, gestational diabetes, maternal depression, antepartum and postpartum hemorrhage, abortion, unwanted pregnancies, and maternal mortality. Additionally, child health outcomes will be assessed, including the prevalence of low birth weight (<2500 grams), stunting (WHO Length-for-Age Z-score <2 SD at one year of age), stillbirth rate (SBR), neonatal mortality rate (NMR), and infant mortality rate (IMR). Sub-clinical parameters for anemia, such as serum ferritin and serum vitamin B12 levels, will also be measured.

## 2. Objective

### 2.1 Primary Objectives

To evaluate the effect of delivering village level PCC through trained ANMs on pre-conception health status among *eligible women* and maternal and child health outcomes in rural Wardha, Maharashtra, India.

### 2.2 Secondary Objectives

To evaluate the factors facilitating and inhibiting the implementation and utilization of the PCC package, the feasibility of the intervention using the RE-AIM framework, and the unit cost of implementing community-based PCC interventions.

## 3. Methods

The conceptual framework and pathways for improving maternal and child health outcomes are grounded in a Theory of Change that illustrates the causal link between targeted interventions and their effect on health. (Figure S1- Supplementary material) illustrates the Pathways and Theory of Change, showing how participatory planning and targeted interventions in PCC improve maternal health, reduce mortality, and promote healthier child outcomes through behavior change.

### 3.1 Study Design

The study will employ an open cohort stepped-wedge cluster randomized design (swRCT) with repeated cross-sectional sampling to measure outcomes. This approach will allow for systematic and phased implementation of the intervention, with gradual exposure of the study clusters to the PCC package, ensuring a robust evaluation of its effectiveness.

#### 3.2.1 Study Setting

The study will be conducted in the villages of the Wardha Community Development block, which are served by four PHCs. These rural settings will provide the context for assessing the feasibility and effect of delivering PCC at the community level.

#### 3.2.2 Study Organization

The trial will be integrated with the District Health System, using the Village Health Nutrition and Sanitation Committee (VHNSC) to promote the PCC package (Figure S2- Supplementary material). Following participatory principles, registered beneficiaries will receive PCC interventions until conception, then transition to standard antenatal, intranatal, and postnatal care through Primary Health Centers (PHCs). Trial staff will ensure continuity of care, following each beneficiary until their child turns one, provided there is a live birth. A Technical Advisory Group (TAG) will oversee the project, offering technical guidance, mentorship, and quality assurance. The Dr. Sushila Nayar School of Public Health (DSNSPH) at Mahatma Gandhi Institute of Medical Sciences (MGIMS), Sevagram, will serve as the social franchiser, supporting VHNSCs in implementing the PCC package in study villages. Under India’s National Health Mission (NHM), VHNSCs are critical in engaging local communities for decentralized health planning. These subcommittees of the village council (Gram Panchayat) drive local health action, raise community awareness, increase healthcare access, and support village healthcare providers. (Ministry of Health and Family Welfare. Government of India, 2013; Ministry of Health and Family Welfare. Government of India., 2015) In the proposed study, based on the earlier experience of the Team at DSNSPH, VHNSC will be catalyzed to act as the *Social Franchisees* for making the PCC services available within their own village. (Pyle et al., 2009)

DSNSPH and participating villages will enter a Social Franchising agreement (Annexure 1- Supplementary material)), signed by the Gram Panchayat, which delegates the responsibility to the VHNSC. As Social Franchisees, VHNSC will organize and guarantee PCC services in the village on behalf of the Gram Panchayat. They will set up and manage pre-conception clinics, providing space, infrastructure, volunteers, and logistical support for operations. Additionally, the VHNSC will promote the PCC package, register beneficiaries, ensure compliance, pay village volunteers, and maintain accountability for the clinics. The PCC package will not be free; beneficiaries will pay a nominal fee to access services, fostering a sense of value for the care provided. The cost of registration, capped at 100/- INR for the entire study duration, will be determined by mutual consensus between potential beneficiaries and the VHNSC. DSNSPH will serve as the Social Franchiser without charging the VHNSC but will regulate the process to ensure the registration fee remains nominal. VHNSCs will have the right to waive the fee for vulnerable individuals or couples unable to pay. Under the Social Franchisee agreement, DSNSPH and VHNSC will collaborate to deliver the PCC package, utilizing existing health system opportunities and related sectors, with the overarching goal of improving maternal and child health outcomes in the village.

### 3.3 Trial design

This study will utilize an open cohort, stepped-wedge cluster randomized design with repeated cross-sectional sampling for outcome measurement, as shown in Figure 1. The study will span approximately three years. All villages will initially be assigned to the control arm, with each village crossing over to the intervention arm at a designated point through randomization. A six-month pre-rollout phase will involve formative research to assess community perceptions and needs, particularly those of potential beneficiaries. This will be followed by a six-month post-rollout phase after all villages have been exposed to the intervention. During the pre-rollout phase, Participatory Learning and Action (PLA) activities will be conducted with the VHNSC, potential beneficiaries, and key stakeholders to identify and finalize the core components of the PCC package through a consultative process. Initially, no individuals will be exposed to the intervention. Gradual exposure will occur as villages transition to the intervention arm, with all villages crossing over by the end of the intervention phase. Crossover will occur in four sequences, each involving 20 villages, with cross-sectional surveys conducted before each crossover using the Probability Proportional to Size (PPS) method to sample participants. (World Health Organization, n.d.) In all there will be 5 timepoints in the study with a cross-sectional survey being conducted at each time point namely at around baseline, 12-months, 18-months, 24-months and 36-months respectively. In each cross-sectional survey, 20 clusters will be selected from each sequence i.e. a total of 80 clusters every cross-sectional survey. A baseline survey will be conducted prior to the Cross-over point 1. The endline survey will happen at Timepoint 5 (36 months). Each cross-over will happen after an interval of 6 months each from the start of trial. Duration of exposure for individuals in Sequence 1, Sequence 2, Sequence 3 and Sequence 4 will be 2.5 years, 2 years, 1.5 years and 1 year respectively.

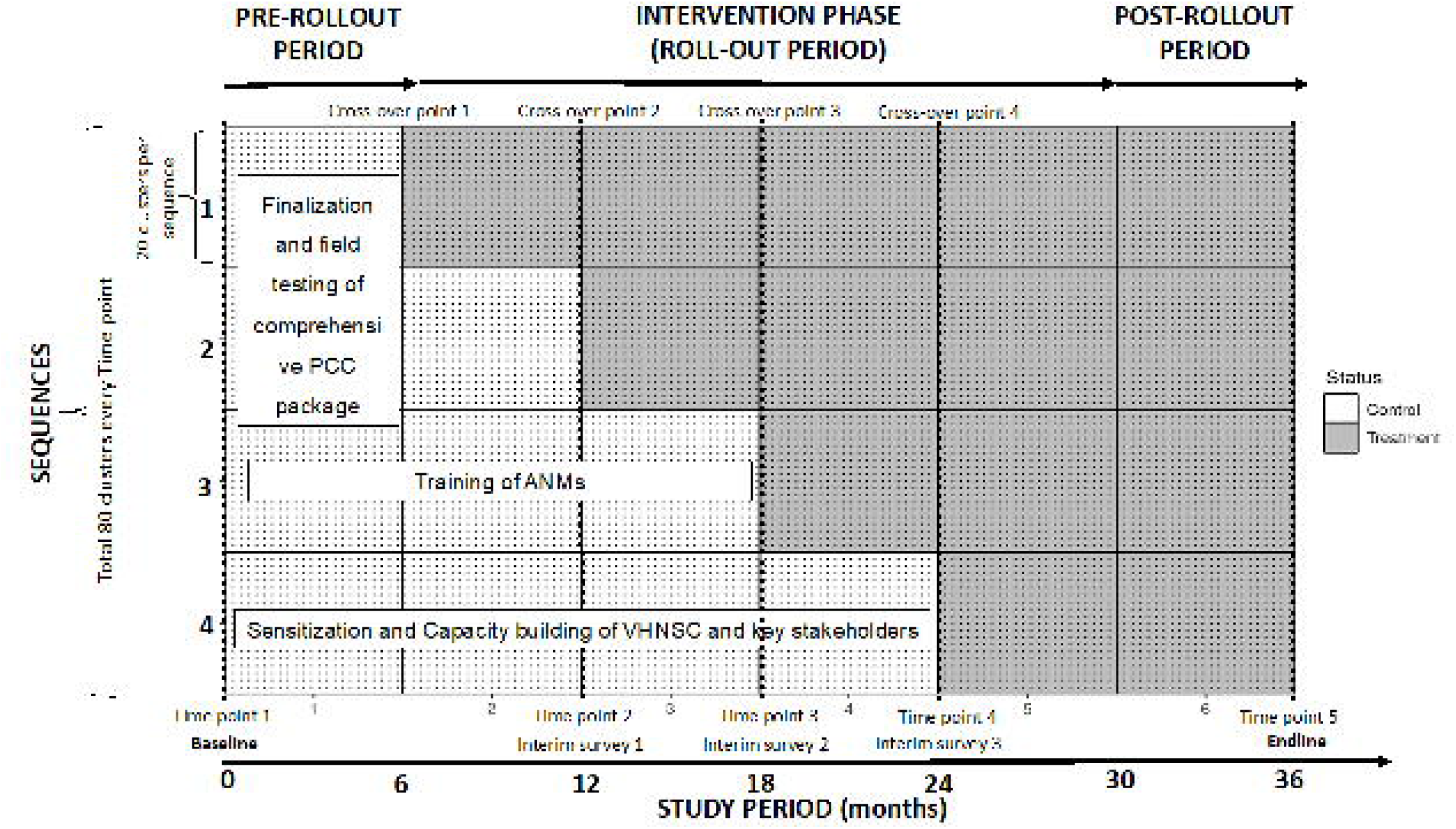

#### 3.4.1 Study Participants, Village Recruitment & Randomization

The study area consists of approximately 96 villages with a total population of around 120,000. The villages vary in size, with populations ranging from 300 to 10,000. According to the 2011 India census, Wardha district had a population of 1,296,157, with males constituting 52% and females 48%. The district experienced a population growth rate of 4.8% between 2001 and 2011 and has a sex ratio of 946 females per 1,000 males. Wardha boasts a literacy rate of 87.2%, which is higher than the national average (64.5%), with male literacy at 92% and female literacy at 81%. Approximately 11.4% of the population lives below the poverty line, and around 73% of the people reside in rural areas, primarily dependent on agriculture. Rural areas are almost entirely reliant on government public health services through PHCs, while private practitioners mainly serve semi-urban and urban areas. (DIRECTORATE OF CENSUS OPERATIONS MAHARASHTRA, 2011)

As shown in Figure 2, villages will be stratified into four categories based on population size: less than 1000, 1000–3000, 3000–5000, and over 5000. Randomization within each stratum will allocate one-fourth of the villages to the intervention arm at each crossover point. A computer-generated randomization list, managed by an independent Data Manager, will be used for assignment. Villages will be randomly assigned to one of four sequences (A to D) until their crossover from control to intervention. The randomization process will remain concealed from investigators and clusters until one month before crossover, allowing time to sensitize VHNSC members and sign Social Franchisee agreements. Blinding of investigators, ANMs, and beneficiaries is not feasible, but survey enumerators and statisticians will be blinded to group allocation to reduce bias.

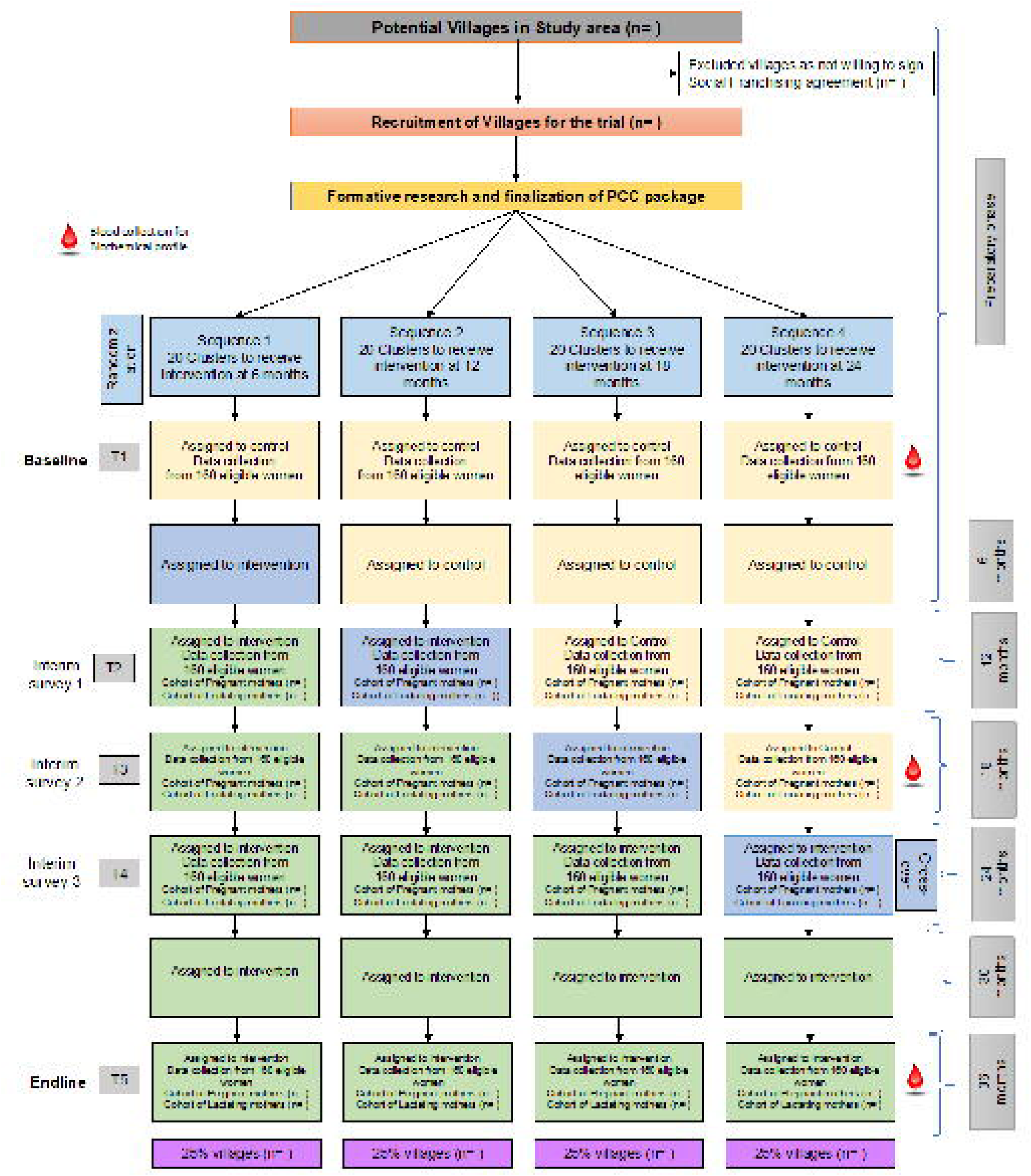

The eligibility criteria for clusters require that all villages under the four PHCs of the Wardha Community Development Block are eligible to participate in the trial. However, only those villages that agree to participate by signing the Social Franchisee agreement will be included, while peri-urban villages within a 5 km radius of Wardha town will be excluded. While, eligibility for including individuals requires that potential participants include women aged 19 to 45 years who have not yet completed their families and are permanent residents of the eligible villages. To be included in the trial, participants must provide written informed consent and be willing to reside in the study area for the next two years. However, couples where either partner has adopted a permanent method of contraception will be excluded from the study.

#### 3.4.2 Operational definitions

- Eligible couple for study purpose will include couples who are newly married or having ≤ 2 children or with an intent to expand the family further wherein wife is in the age group of 15 to 45 years. (National Health Mission., 2013)
- Pre-conception period for study purpose will be considered as the time point from when individual/couples first decide to become pregnant till they conceive. It will include the 3 months before conception as per the standard definition or extend beyond that. (Stephenson et al., 2018)
- Contraceptive prevalence is the percentage of women or whose sexual partner are currently using, at least one modern method of contraception, regardless of the method used. (World Health Organization, 2016)

#### 3.4.3 Current Gaps in Preconception Care Delivery

India currently lacks a dedicated public health program for regular PCC services, including in the study area. PCC is typically offered through private clinicians, health facilities, or research settings evaluating interventions, and only those who opt-in or participate receive care. While the WHO recommends 13 areas for PCC, some are addressed by national health programs. For example, the RMNCH+A program promotes early pregnancy registration (by 12 weeks) for ANC and provides Iron Folic Acid (IFA) supplements to adolescent girls and women of reproductive age. Contraceptive options are also available under RMNCH+A, and a Measles-Rubella vaccination campaign targets girls under 15 years, though it is not part of routine immunization for older adolescents or married women who missed rubella vaccination. Other programs, like those for tobacco control and genetic screening under RBSK, exist but face challenges with coverage and implementation, as indicated by National Family Health Survey-4 (NFHS-4) data. (INTERNATIONAL INSTITUTE FOR POPULATION SCIENCES, 2016; International Institute for Population Sciences, 2016) Under the NHM, Village Health and Nutrition Days (VHND) are organized monthly in each village. Accredited Social Health Activists (ASHAs) and Anganwadi Workers (AWWs) mobilize women and children to gather at the Anganwadi Centre, where ANMs and PHC staff provide antenatal, postnatal care, immunization, contraceptive services, growth monitoring, newborn care, and health counseling. The VHNSC and Gram Panchayat assist in organizing these events. (Ministry of Health and Family Welfare & Government of India, n.d.) During the control period, the population will continue to receive the routine services and care through the existing government programs and no additional activities would be conducted during the control period.

### 3.5 Sample size and power analysis

A sample size of 3,200 (640 at each of five time points) was estimated using the R software package for a swRCT, employing a Gaussian distribution and accounting for discrete time decay. (Hakhu et al., 2019; Hemming et al., 2020) The assumptions used were a two-sided statistical significance level of 95%, alpha = 0.05, four sequences, 20 clusters per sequence, power of 80%, eight observations per cluster at each time point, effect size of 0.2, standard deviation (σ) of 2.0, intra-cluster correlation (ICC) of 0.2, cluster auto-correlation (CAC) of 0.8, and a fractional treatment effect of 0.5. An assumed improvement of 20% in the proportion of women from eligible couples entering pregnancy in good health (without undernutrition, anemia, or depression) is considered the minimum meaningful public health effect. (Figure S3- Supplementary Material)

For secondary maternal and child health (MCH) objectives, a cohort of potential beneficiaries will be followed from conception through pregnancy outcomes and until the child turns one year old for live births. A similar number of individuals from control areas will be recruited for comparison. Biochemical profiling will be conducted on a subsample of 1,920 individuals (640 each) at baseline, an interim survey 3 at 18 months, and endline. Barrier analysis will be undertaken for assessment of the facilitating and inhibiting factors for implementation and utilization of PCC package in the last year of project. (Kittle, 2013) The sample size provides 90% power to detect a 1-unit difference in mean outcomes for continuous variables. This difference, considered the minimum meaningful clinical change, would make the intervention relevant, such as improving hemoglobin levels by at least 1 g/dL in participants.

### 3.6 Implementation strategy

#### 3.6.1 Pre-conception care package Intervention

The intervention involves delivering a comprehensive PCC package through village-based clinics led by trained ANMs. The package includes evidence-based interventions and health promotion messages on nutrition, tobacco and alcohol use, contraception, reproductive health, responsive caregiving, vaccination, genetic conditions, environmental health, infertility, interpersonal violence, and mental health, as recommended by WHO. (World Health Organization, 2013) The proposed PCC clinics will run on the same day in parallel to the VHND in the village. (Ministry of Health and Family Welfare & Government of India, n.d.) The existing PHC ANMs will provide services during VHNDs, while project-specific ANMs, recruited and trained for the study, will deliver PCC services at village clinics to complement the government’s services. These ANMs will be closely monitored by the investigators for intervention fidelity. VHNSCs will provide space for the PCC clinics, and all registered beneficiaries will receive the comprehensive PCC package.

The PCC package, including its contents, interventions, delivery, and number of visits, will be finalized through a consultative process with potential beneficiaries to ensure services meet their needs. The package will feature evidence-based interventions promoting health in key areas: Nutrition (BMI, hemoglobin), Tobacco and Alcohol use (addiction assessment), Contraception (use assessment, suggestions), Reproductive health (STI/RTI screening, referral), Responsive caregiving (feeding, age-appropriate stimulation), Vaccination (Rubella, Hepatitis B, tetanus, diphtheria), Genetic conditions (family history, referral), Environmental health (hazard exposure, counseling), Infertility (assessment, referral), Interpersonal violence (screening, referral), Mental health (PHQ-4, referral), Physical activity (assessment, promotion), and Stress mediation (stretching, meditation, breathing exercises). This comprehensive package holistically addresses women’s health needs during the preconception period. Evidence-based strategies will be finalized through participatory planning with stakeholders, allowing beneficiaries to identify initiatives that are both acceptable and feasible, such as promoting iron-rich food, IFA use, contraception, stress mediation, and physical activity. Community mobilization will promote the value of PCC services, encouraging their use. Every six months, community events will be organized by VHNSC with support from frontline health workers. DNSNPH and trial staff will provide technical assistance, and all stakeholders can participate freely. VHNSC will manage these events under the Social Franchisee agreement.

#### 3.6.2 Preparatory Phase

Preparatory phase will be used for liaising with concerned District authorities from health, panchayat raj and other relevant departments. Project staff recruitment will also be done in the first month of preparatory phase.

#### 3.6.3 Formative research for understanding stakeholder perspectives around PCC

In-depth/ key informant interviews, focus group discussions (FGDs), free-listing, and ranking exercises will be used to understand community perceptions of PCC. Findings from the formative study will shape the final PCC package, central to its implementation in the roll-out phase. The package will use an Incremental Learning Approach, focusing on reaching every potential beneficiary and tailoring delivery mechanisms. The participatory nature of the package enables stakeholders to implement contextually appropriate initiatives, such as stress mediation strategies and increasing iron-rich food consumption as shown in Figure 3.

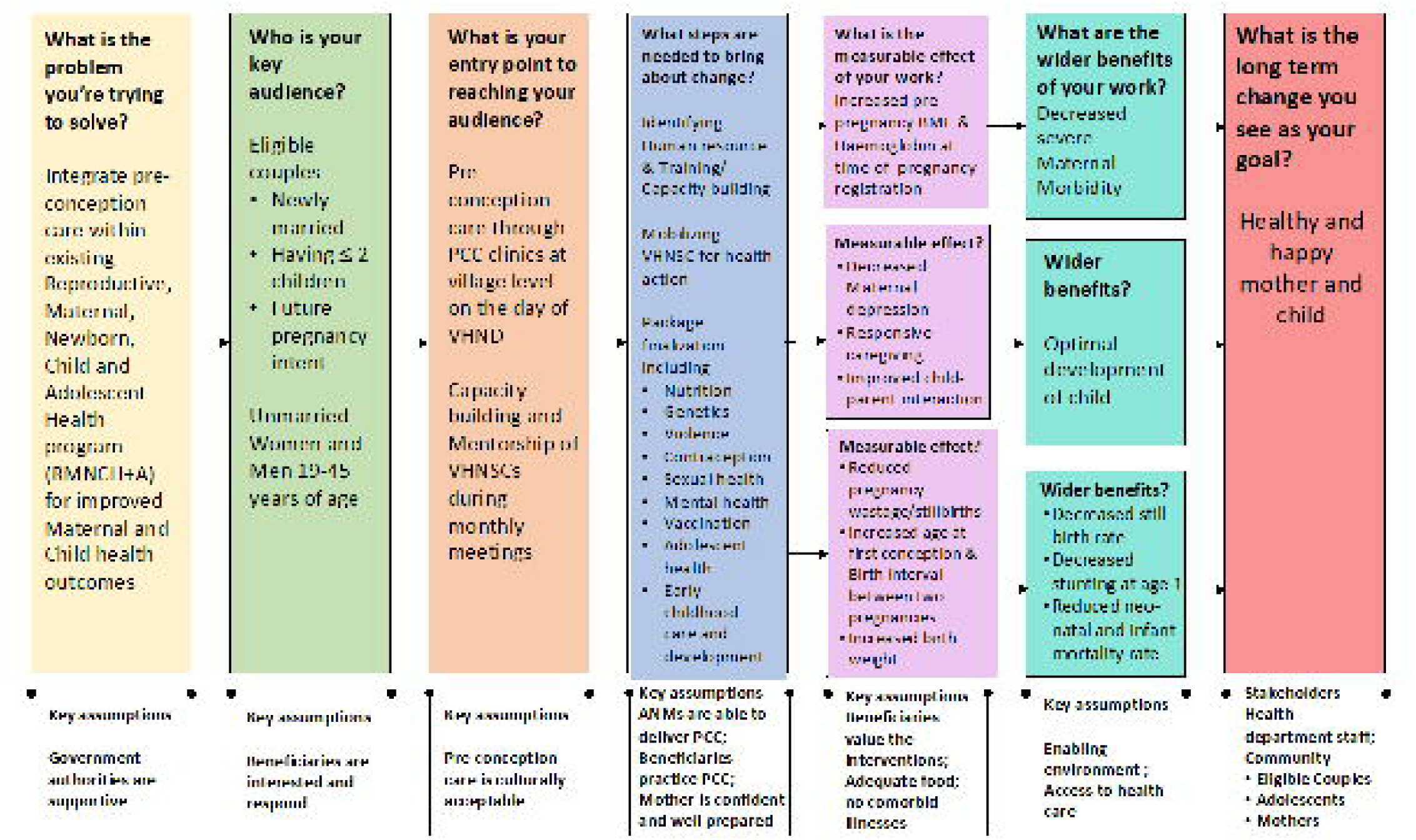

#### 3.6.4 Technical field testing

The comprehensive PCC package will be field-tested in select villages of a neighboring PHC before implementation in the study area. This testing, conducted as planned for the main trial, will help refine content and strategies. Data from field testing will be kept separate from the main trial data.

### 3.7 Data collection

The Data Collection Tool (DCT), aligned with study objectives, will be pre-tested and used on handheld tablets via Open Data Kit, capturing individual surveys, anthropometry, and hemoglobin levels. It will gather baseline demographic details and health behaviors like IFA consumption, diet, physical activity, stress mediation, interpersonal violence, tobacco, alcohol use, contraceptive use, pregnancy intent, and caregiving. The DCT will also record vital events (births, stillbirths, abortions, deaths) and psychological assessments (PHQ-4), along with physical measurements (height, weight, birth weight) and biochemical assessments (hemoglobin). It will track PCC registration, intervention compliance, PCC visits, and adverse effects from interventions. Procedures will follow NFHS and WHO-STEP guidelines.(International Institute for Population Sciences, 2014) Trained ANMs will record all readings to minimize inter-observer variation, with training provided to avoid digit preference and expectation errors. Hemoglobin estimation will involve collecting 2 ml venous blood samples, which will be promptly transferred to the central laboratory. Participants’ identities will remain confidential, with unique identification numbers assigned, and personal details will not be disclosed, including in scientific publications. Data quality will be thoroughly checked before analysis.

### 3.8 Statistical Data analysis

Data analysis will follow the Intention to Treat (ITT) principle, comparing outcomes between control and intervention groups, with villages analyzed according to their randomized crossover time, regardless of whether crossover occurred as planned. An “As Treated” analysis will also be conducted based on actual allocation. Baseline characteristics will be presented for the entire study and stratified by cluster and group allocation. The intervention’s effect will be assessed using Generalised Linear Mixed (GLM) regression models, adjusting for time and within-cluster correlations. Interim analysis will be conducted after each cross-sectional survey. R software will be used for statistical analyses, and WHO Anthro Survey software will analyze infant anthropometry data.

*Additional analysis:* A subgroup analysis will compare outcomes between those registered for PCC services and non-registrants. Among registrants, outcomes will be analyzed based on intervention compliance. Additional analyses will examine factors affecting compliance and extended effects on caregiving, pregnancy wastage, and health behaviors. Sensitivity and post-hoc power analyses will assess robustness, with post-hoc analyses clearly identified.

#### 3.9.1 Quality monitoring

Standard operating procedures (SOPs) will be developed for anthropometry, including measuring weight and length/height using calibrated electronic scales and stadiometers. SOPs will also cover blood collection and sample transport for hemoglobin estimation, performed in the institute’s accredited central laboratory. Investigators will randomly backcheck 1% of forms to ensure data quality. Supportive supervision checklists will monitor PCC session quality and intervention fidelity (Figure S1) & (Figure 3).

#### 3.9.2 Safety and data monitoring

As a health promotion trial, it poses minimal risk to participants with few safety concerns. The Technical Advisory Group (TAG) will review interim data every six months, ensuring inclusivity and monitoring adverse events, stillbirths, abortions, and deaths to ensure they remain within expected limits.

## 4. Discussion

The proposed project is an attempt for implementation of PCC interventions at village level in *real* life-settings and will help to develop a PCC package that could be delivered through a task-sharing approach by the trained ANMs. The proposed trial will be first of its kind in India for implementation of PCC interventions at village level. The trial will help to find out the *felt need* and acceptance for pre-conception care in a rural area of India. The study will help to bridge the gap in existing strategy that begins with early registration of pregnancy.

The trial will be a value addition to the existing public health programs as it will assess the feasibility of implementing a community-based package for PCC complementary to the existing programs. The proposed research has significant potential to improve population health by reducing adverse pregnancy outcomes such as abortion, severe maternal morbidities, stillbirth, low birth weight, and early-neonatal mortality. It will also contribute to lowering infant mortality rates (IMR) and stunting at age one. Additionally, the study will assess the impact of preconception interventions on sub-clinical anemia, potentially advocating for evidence-based treatment instead of blanket IFA supplementation. Barrier analysis will identify challenges and facilitators for implementing the PCC package in rural areas, refining the approach for broader application. If effective, this PCC model could enhance India’s RMNCH+A program, maximizing maternal and child health outcomes and providing a valuable addition to existing strategies.

### 4.1 Expected Outcome

The primary outcome of the study will be the improvement in pre-conception health among women from eligible couples, measured by the proportion of women entering pregnancy in good health, free from undernutrition, anemia, and depression as shown in Table 1 and Figure 3. The secondary outcomes will include improved maternal and child health, such as reductions in stillbirth rate, neonatal mortality rate (NMR), infant mortality rate (IMR), and severe maternal morbidities (e.g., pre-eclampsia, gestational diabetes, maternal depression, hemorrhage, and abortion). Other outcomes include lower rates of low birth weight, stunting at age one, delayed first pregnancies, longer birth intervals, and more planned pregnancies. To assess feasibility, the RE-AIM framework (Table 2) will be used to evaluate the intervention’s feasibility, acceptability, and effectiveness. Additionally, the unit cost of implementing community-based PCC interventions will be measured, accounting for direct, indirect, and opportunity costs, aiding future policy advocacy for the PCC package.

**Table 1:**
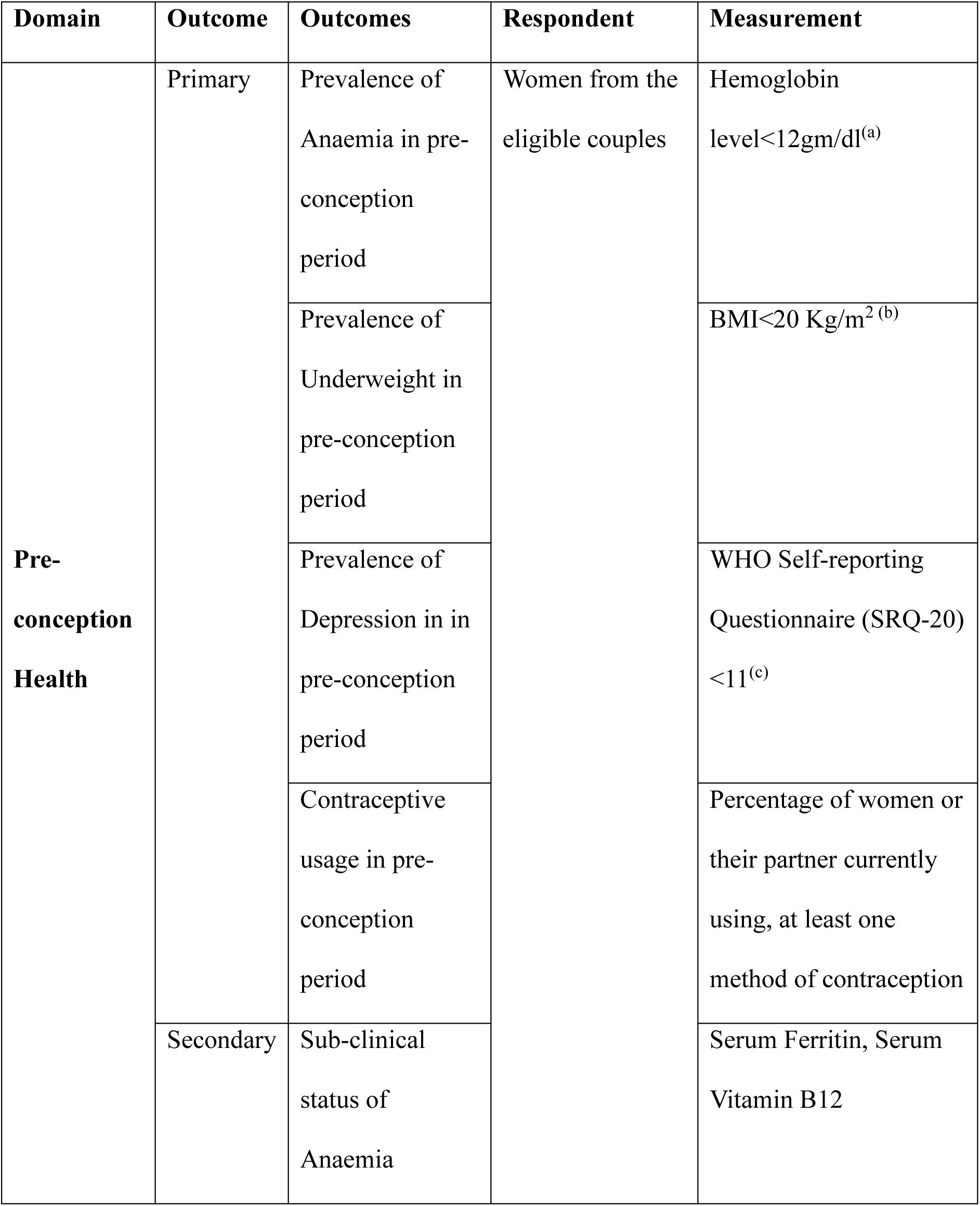
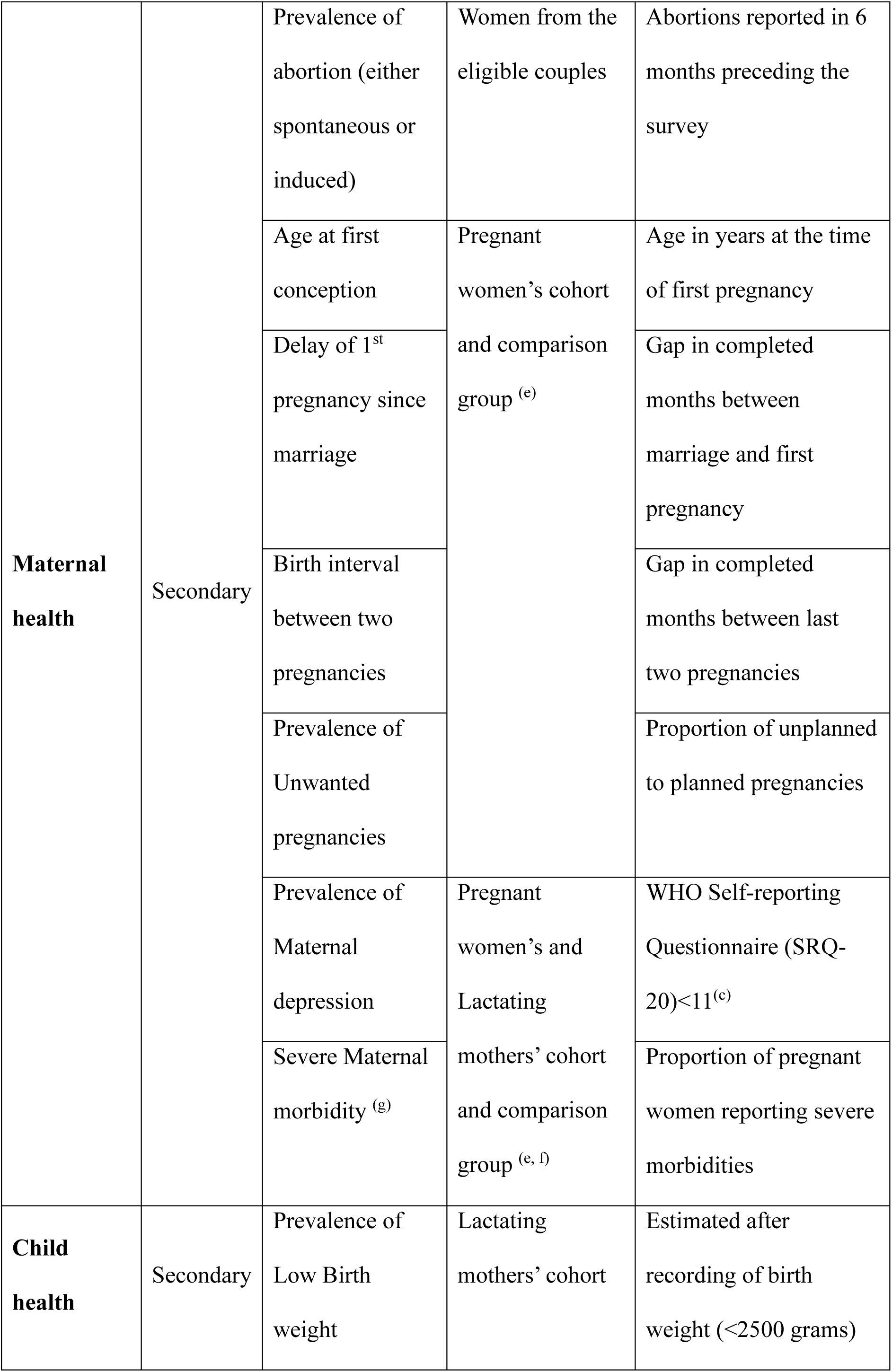
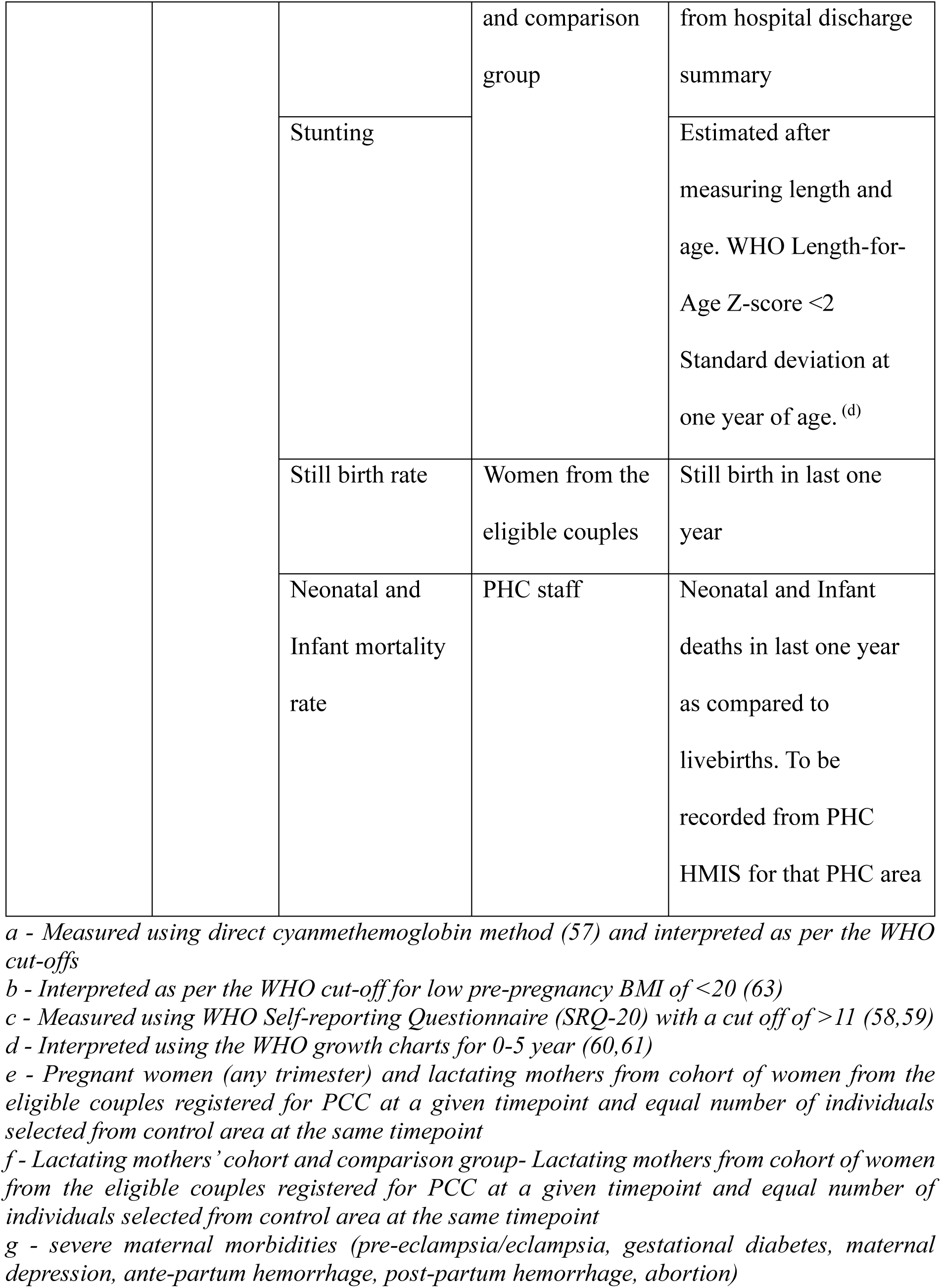
Outcome Measures for Pre-Conception, Maternal, and Child Health in a Community-Based Pre-Conception Care Intervention.

**Table 2:** Framework to assess the feasibility of intervention using the RE-AIM framework.

| **Domains** | **Questions** | **Measures (quantitative/qualitative)** |
| --- | --- | --- |
| **Reach** | What is the enrollment, retention, and follow-up rates of the participants through the study and intervention?<br>What are the reasons for adherence rates to intervention, intervention attendance, and engagement? | • Characteristics of participants<br>• Proportion of participants enrolled for the study<br>• Proportion of participants completed all the visits<br>• Proportion of participants adhered to the advices<br>• Proportion of women in good health<br>○ without under nutrition<br>○ without anemia<br>○ without depression<br>• improvement in MCH outcomes<br>• Proportion of drop-out participants<br>• Satisfaction with the delivery of the PCC package (component wise) |
| **Adoption** | What was the acceptability and suitability of the intervention | • Number of visits attended<br>• Proportion of participants opted for healthy lifestyle (changed dietary habits, involved in physical activity)<br>• Proportion of participants advised vaccination got vaccinated |
|  |  | • Proportion of participants advised IFA supplementation, consumed IFA tablets<br>• Participant perception of intervention<br>• Challenges and facilitators during the implementation |
| **Effectiveness** | Is there evidence that the intervention produced a change in the desired outcomes<br>Utilization and quality of delivery of the intervention | • Change in baseline and end-line characteristics<br>• improved preconception health status (delayed pregnancy, contraceptive practice, anemia, underweight, depression)<br>• improved maternal outcomes (birth interval, unintended pregnancy, depression, severe maternal morbidity)<br>• improved child outcomes (decreased still birth rate, neonatal and infant mortality rate)<br>• Participant perception<br>• Providers perception |
| **Implementation** | What was the uptake and utilization of the intervention | • Proportion of participants completed the intervention<br>• Proportion of participants in good health<br>• feasibility of the recruitment process |
|  |  | • Overall satisfaction of participants with the intervention |
| **Maintenance** | Are desired health behaviors sustained | • The proportion of married women delaying conception<br>• Proportion of participants willing to spread the message of pre-conception care |

Future strategies will be implemented at the community level through local gatherings and meetings. A communication and capacity-building plan will be developed in the initial phase, closely tied to the research findings. If proven effective, advocacy will be carried out at the Maharashtra state and national levels to integrate the PCC package into existing programs, enhancing their impact.

### 4.2 Strengths & Limitations of the research

The project’s alignment with the District Health System and integration into the VHNSC structure enhances community acceptance and long-term sustainability. By leveraging *participatory development principles*, local stakeholders are empowered, fostering true community ownership. The evidence-based PCC package is tailored to the community, ensuring acceptability and feasibility, increasing the likelihood of meaningful impact on maternal and child health. External generalizability of the study will be limited to other rural areas with similar profile and not for the urban or tribal areas.

## Data Availability

The datasets that will be used and/or analyzed during the study will be available from the corresponding author upon reasonable request.

## Ethics approval

Ethics approval was obtained from the Institutional Ethics Committee (IEC) for Research on Human subjects at MGIMS, Sevagram, Maharashtra, India before the trial initiation, as per IEC approval vide letter no (IEC/406/2023, dated 30 December 2023). Written informed consent will be taken from all adults before enrolment. Participants diagnosed with health issues during PCC visits will receive treatment at the Primary Health Centre, with referrals provided if necessary. Interviews and examinations will be conducted privately, with a female present during women’s examinations. All study records will remain strictly confidential.

## Abbreviations

PCC: Pre-conception care
ANM: Auxiliary Nurse Midwives
VHNSC: Village Health Nutrition and Sanitation Committee
VHND: Village Health Nutrition Day
RMNCH+A: Reproductive, Maternal, Newborn Child and Adolescent Health
NMR: Neonatal mortality rate
IMR: Infant mortality rate
GLM: Generalised Linear Mixed
swCRT: stepped wedge cluster Randomized Controlled Trial
SDG: Sustainable development goals
PHC: Primary Health Centre

