## Supplementary_Material.pdf for "Evaluating the effects of a community-based Pre-Conception Care package through trained nurses on Pre-conception Health, Maternal and Child Health Outcomes: Protocol of a Stepped-wedge Cluster Randomized Implementation Trial"

**Figure S1: Pathways and Theory of Change for the effect of interventions to improve maternal and child health outcomes**

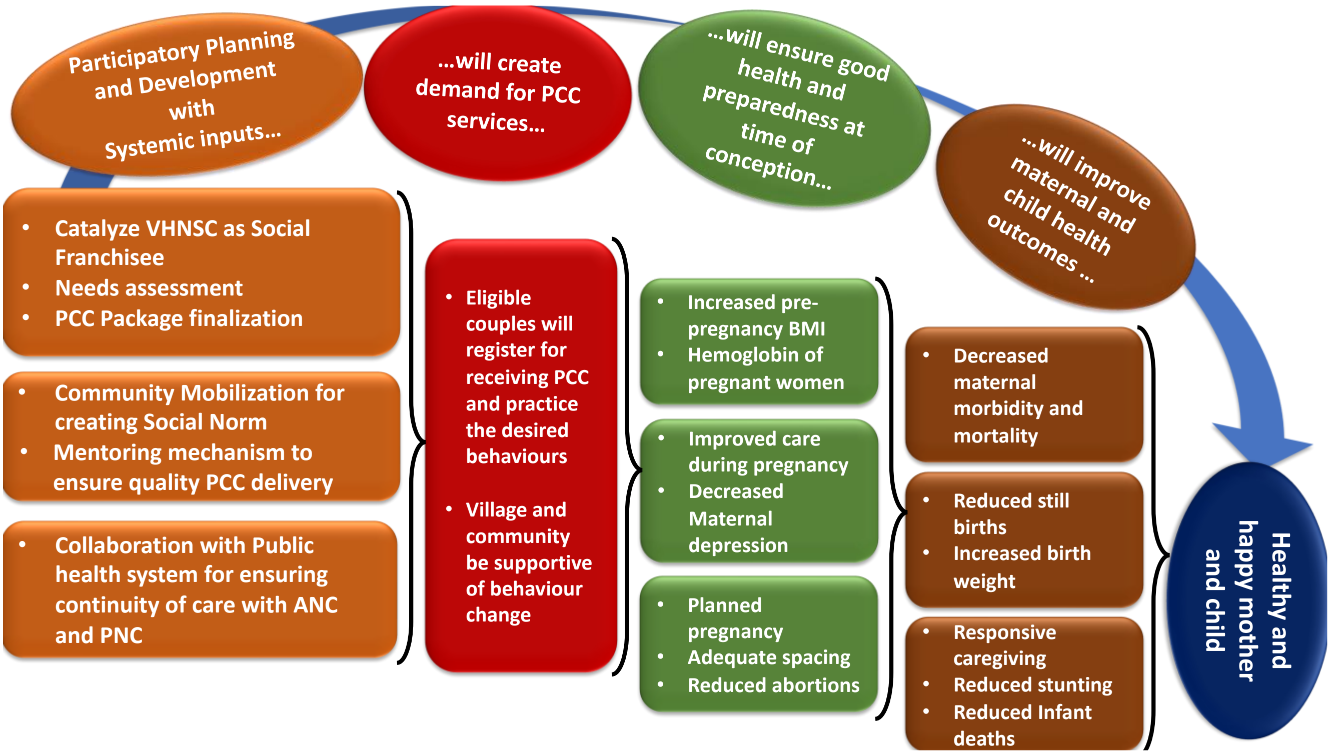

**Figure S2: Framework for Implementation of the Pre-Conception Care Package Through Social Franchising and Village Health Systems**

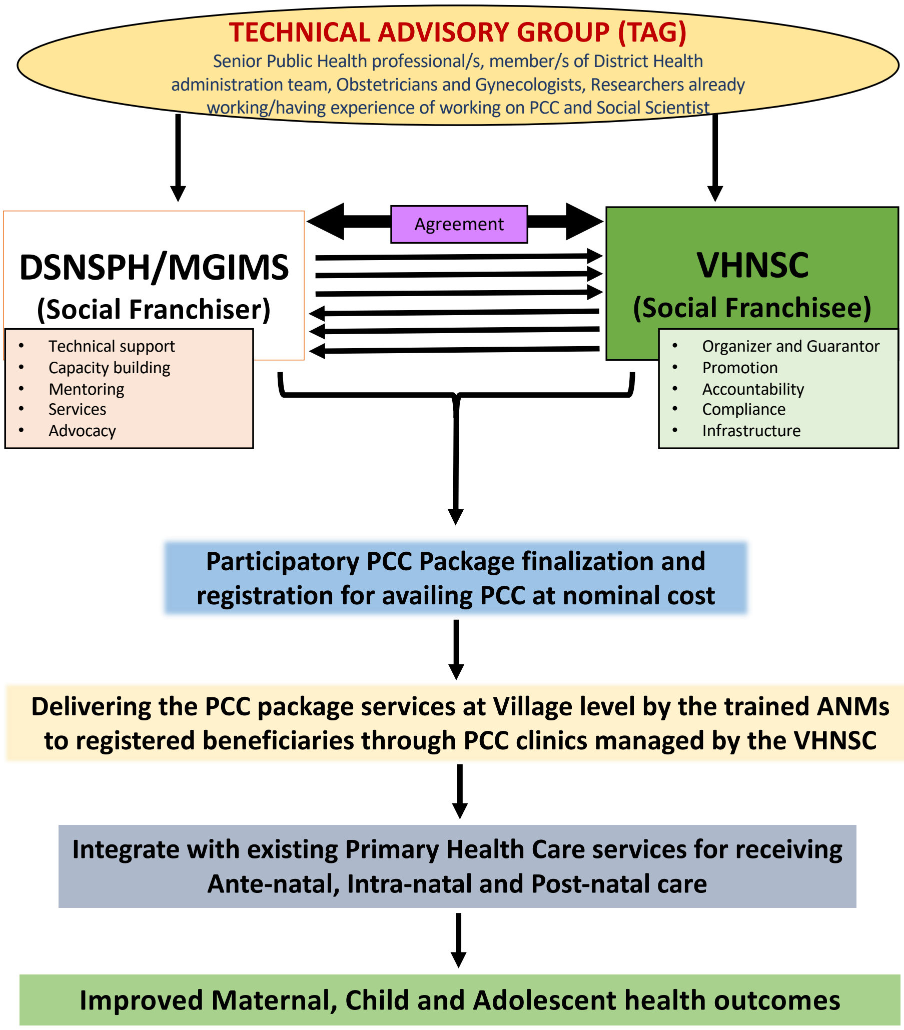


- PCC - Preconception care
- VHNSC - Village Health Nutrition and Sanitation Committee
- VHND - Village Health Nutrition Day
- DSNSPH - Dr Sushila Nayar School of Public Health
- MGIMS - Mahatma Gandhi Institute of Medical Sciences

**Figure S3: Power Curves for Different Intra-cluster Correlation Coefficients (ICC) and Cluster Sizes in a Stepped-Wedge Cluster Randomized Trial**

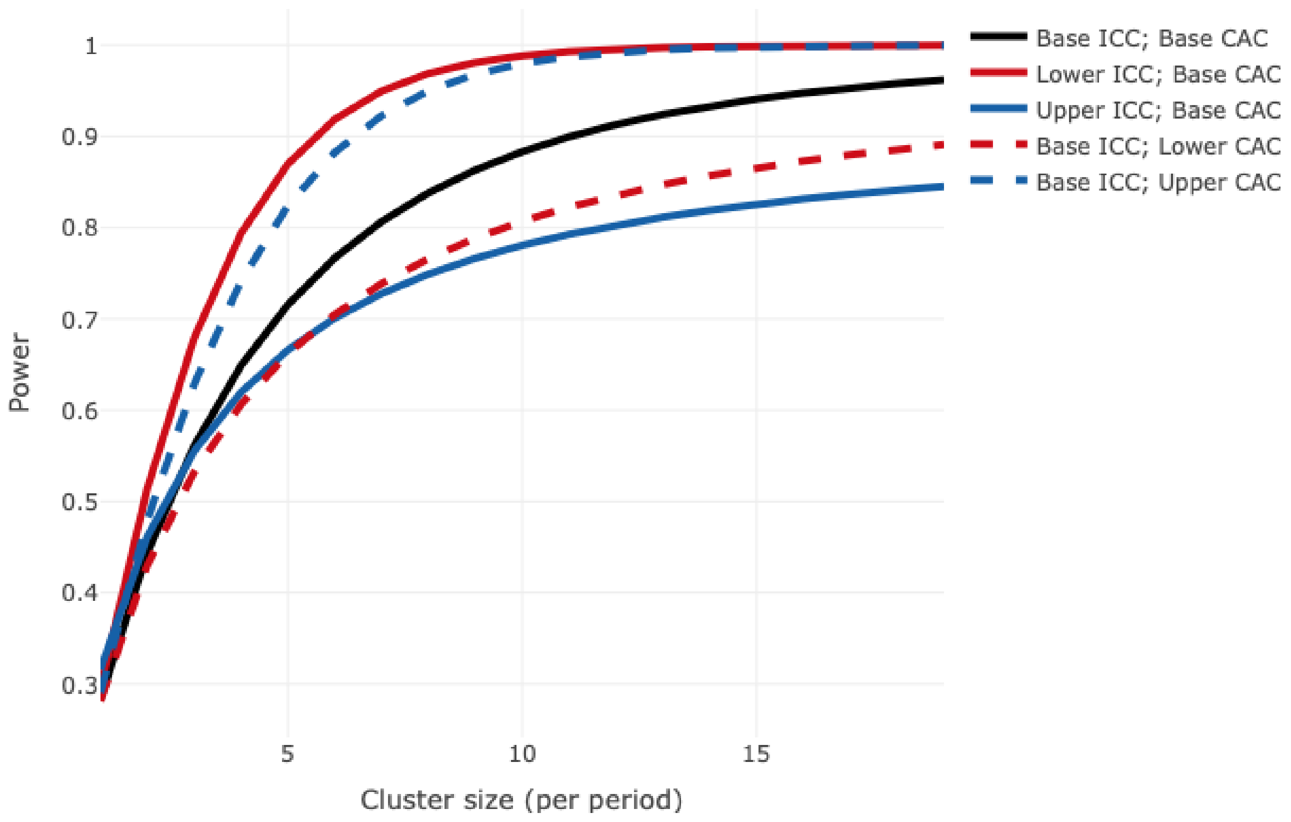

**Annexure 1: Social Franchisee Agreement (Draft)**

The Village Health Nutrition and Sanitation Committee (VHNSC) (promoted under the National Health Mission to act as leadership platforms for *"local level community health action"* representing the Gram/Village Panchayat) of village----------under Primary Health Center (PHC)-----------(hereinafter called the 'Social Franchisee') hereby enters into an agreement with the Dr Sushila Nayar School of Public Health (DSNSPH), Mahatma Gandhi Institute of Medical Sciences (MGIMS), Sevagram, Wardha, (hereinafter called the 'Social Franchiser') for planning and managing comprehensive Pre-conception Care (PCC) clinics at the village-level for improving maternal, newborn and child health outcomes (hereinafter referred to as the Social Product) in accordance with the following terms and conditions:

**The Franchisee must:**

- Function as a decision-making body to manage PCC clinics in their village
- Conduct a community health needs assessment and engage in participatory program planning
- Be able to plan a Social marketing strategy and generate demand for PCC services at village level
- Have in place an operational mechanism to promote the PCC package within the village.
- Decide the cost of PCC voucher mutually through consensus with the potential beneficiaries
- Have in place an operational monthly information system for review of work done and sharing of information with Frontline Healthcare workers and DSNSPH project staff.
- Ensure that the Social Product reach at least of 60% of the population
- Have in place a well-defined and controlled cost recovery mechanism for the PCC vouchers
- Be able to understand the respective roles of Franchisee and Franchiser.
- Be willing to accept the Franchiser as a technical support and monitoring agency
- Be willing to waive off the charges for PCC vouchers for poor and/or vulnerable individuals/couples who do not have the capacity to pay for the vouchers.

**Have the ability to implement and manage the following:**

1. Provide Space and Infrastructure for PCC clinics
2. Nominate and pay honorarium to village volunteers for organizing PCC clinics
3. Promotion of PCC package within the village.
4. Social marketing of PCC vouchers to potential beneficiaries
5. Registration of beneficiaries for availing PCC services
6. Ensuring compliance of registered beneficiaries
7. Printing, maintenance and accountability of PCC vouchers
8. Referral linkages with public and private health care facilities
9. Organize Community based events for creation of Social norm for availing PCC

**The Franchiser must:**

- Provide the necessary technical support to PCC clinics
- Quality improvement of PCC services through PCC clinics at village level
- Build the capacity of the VHNSC to conduct a community health needs assessment, plan and implement PCC interventions, develop an information system, communications strategy, alternative financing for PCC vouchers and sustainability plan.
- Mentorship of VHNSC for sustainability of the intervention

- Assist in selection of village volunteers for PCC clinics
- Develop accreditation guidelines which the VHNSC must meet in order to retain the Social Franchisee status.
  Under the aegis of discharging its Social Accountability, DSNSPH, MGIMS will not charge anything to the VHNSC for being the Social Franchiser, however, it will act as regulator to ensure that the cost of PCC voucher is kept nominal.
  The Franchiser will retain the right to withdraw services in cases where the Franchisee is not following the above terms and conditions.

Signed on -----------------day of----------------------month of-------------------year
By --------------------------------------- ------------ (Name and Address) on behalf of Franchisee
By ----------------------------- (Name and Address) on behalf of the Franchiser.

(Signature) (Signature)
**Franchiser** **Franchisee**

Witness:
--------------------------- (Name and Address)
--------------------------- (Name and Address)
(One of the witnesses should be the village Sarpanch or headman
